# Performance of ChatGPT and Bard in Self-Assessment Questions for Nephrology Board Renewal

**DOI:** 10.1101/2023.06.06.23291070

**Authors:** Ryunosuke Noda, Yuto Izaki, Fumiya Kitano, Jun Komatsu, Daisuke Ichikawa, Yugo Shibagaki

**Author notes:** **Corresponding author** Ryunosuke Noda, MD, The address of all authors: 2-16-1 Sugao, Miyamae-Ku, Kawasaki, Kanagawa 216-8511, Japan.

## Abstract

**Background:** Large language models (LLMs) pretrained on vast amounts of data have significantly influenced recent advances in artificial intelligence. While GPT-4 has demonstrated high performance in general medical examinations, its performance in specialised areas such as nephrology is unclear. This study aimed to compare ChatGPT and Bard and their potential clinical applications in nephrology.

**Methods:** Ninety-nine questions from the Self-Assessment Questions for Nephrology Board Renewal from 2018 to 2022 were presented to two versions of ChatGPT (GPT-3.5 and GPT-4) and Bard. We calculated the overall correct answer rates for the five years, each year, and question categories and checked whether they exceeded the pass criterion. The correct answer rates were compared with those of the nephrology residents.

**Results:** The overall correct answer rates for GPT-3.5, GPT-4, and Bard were 31.3% (31/99), 54.5% (54/99), and 32.3% (32/99), respectively, thus GPT-4 demonstrated significantly higher performance than GPT-3.5 (p < 0.01) and Bard (p < 0.01). GPT-4 met the passing criteria in three years. GPT-4 demonstrated significantly higher performance in problem-solving, clinical, and non-image questions than GPT-3.5 and Bard. The correct answer rate for GPT-4 was intermediate between the rates for third- and fourth-year nephrology residents.

**Conclusions:** GPT-4 significantly outperformed GPT-3.5 and Bard and met the Nephrology Board renewal standards in three of five years. These findings underline the potential applications of LLMs in nephrology as well as their advantages and disadvantages. As LLMs advance, nephrologists must understand their performance and reliability for future applications.

## INTRODUCTION

Rapid advancements in artificial intelligence (AI) have led to high expectations for its applications in the medical field. Among the myriad AI technologies, large language models (LLMs) have demonstrated significant improvements in performance in recent years, which have increased their prominence in AI research [1]. One such application of AI is LLMs, which can generate human-like text and answer prompts based on patterns learned during training. The applicability of LLMs has spread to various fields, including medical education and clinical support, and is expected to provide opportunities for breakthrough advances [2–4]. Comparing the performance of LLMs in medical tasks with that of human experts serves as an important metric for verifying the potential use of AI in medical practice, with particular interest in evaluating the performance of AI in medical examinations [3,4].

ChatGPT is a conversational AI that includes an LLM series known as the Generative Pre-trained Transformer (GPT) series, released by Open AI in November 2022. It responds to multilingual questions in English, Japanese, and other languages on the Internet [5]. In March 2023, a new version of GPT-4 was released, and as of May, two versions, GPT-3.5 and GPT-4, are available for ChatGPT. GPT-4 achieved an approximate accuracy of 90% in the United States Medical Licensing Examination (USMLE) and has consistently met the passing criteria in the Japanese National Medical Licensing Examination for five consecutive years, demonstrating high performance in general medical examinations [6]. The performance of ChatGPT in specialised medical examinations in various fields, such as orthopaedic surgery, gastroenterology, plastic surgery, cardiology, dermatology, and radiology, is also beginning to be reported [7–12].

Bard is a conversational AI released by Google in March 2023. It became compatible with the Japanese input and output in May, and its base LLM changed from LaMDA to PaLM2 [13]. Unlike ChatGPT, which generates an output based on information trained before September 2021, Bard can gather information from the Internet, leading to expectations of its performance in various fields that require more recent information. In terms of medical tasks, Med-PaLM2, a fine-tuned PaLM2 with additional medical datasets, has also been reported recently [14].

However, the performance of conversational AIs, such as ChatGPT and Bard, in the field of nephrology, as well as their differences have not been adequately studied. Furthermore, many aspects regarding the comparison between LLMs and human nephrologists are unknown. The credentials for nephrology expertise are board-certified nephrologists from the Japanese Society of Nephrology [15]. One of the renewal requirements for this certification is a correct answer rate of ≥60% in the Self-Assessment Questions for Nephrology Board Renewal (SAQ-NBR) conducted annually by the society [16]. The SAQ-NBR consists of Japanese written exam questions specialising in nephrology in a multiple-choice format, with five options for each question.

In this study, we evaluated the performance of LLMs in nephrology examinations by comparing the correct answer rates of ChatGPT (GPT-3.5 and GPT-4), Bard, and nephrology residents. We examined the current state and future potential of AI in medical education and its clinical application in nephrology.

## MATERIALS AND METHODS

The SAQ-NBR were answered by ChatGPT GPT-3.5 version (ChatGPT-3.5), ChatGPT GPT-4 version (ChatGPT-4), Bard, and three residents participating in a Japanese nephrology fellowship programme. Correct answer rates were calculated and compared by exam year, overall, and across four categories (taxonomy, question type, image, and subspeciality).

### SAQ-NBR

In this study, we used 99 of 100 questions from the SAQ-NBR from 2018 to 2022, excluding one question deemed inappropriate by the Japanese Society of Nephrology, the issuer of the questions. We classified the questions into four categories: taxonomy, question type, image, and subspeciality. For the taxonomy, we adopted the classification used in the question creation manual for the Japanese National Medical Examination created by the Japan Medical Association, categorising it into recall, interpretation, and problem-solving [17]. Taxonomy refers to the classification based on how much thinking process is required for the examinee to answer questions, with recall, interpretation, and problem solving, in that order, requiring more advanced intellectual abilities. General questions testing knowledge in the field of nephrology and clinical questions regarding specific case management were identified. Questions were classified based on the presence or absence of images, such as renal pathology or computed tomography scans. Subspeciality was based on an experience case list from the Japanese Society of Nephrology: chronic kidney disease /end-stage kidney disease; acute kidney injury; glomerular disease; tubulointerstitial disease; hypertension; renal vascular disease; water, electrolyte, and acid–base abnormalities; autosomal dominant polycystic kidney disease; and urological diseases [18]. This list was supplemented with basic medicine (physiology and pharmacology) for a total of eight items.

### ChatGPT and Bard

The SAQ-NBR questions were input as prompts to ChatGPT-3.5, ChatGPT-4, and Bard, and each was asked to answer. For each year, the prompt “Please answer the following question.” was initially input in Japanese, and then the original text of each question was input individually. Because the current versions of ChatGPT and Bard cannot input images, only the text of the questions was used as input. The correct answer to each question was determined based on the official answers provided by the Japanese Society of Nephrology. ChatGPT-3.5, ChatGPT-4, and Bard were asked to answer similarly, and their correct answer rates were compared. For ChatGPT, ChatGPT Plus, which is available in both GPT-4 and GPT-3.5 versions, was selected. The ChatGPT Plus adopted was the ChatGPT Mar 23 version, released on March 23, 2023, and all prompts were input from April 1 to 5, 2023. For Bard, we used the Japanese-supported β version released on May 11, 2023, and input all prompts on May 15, 2023.

### Residents of a Japanese Nephrology Fellowship Programme

The co-authors Y.I., F.K., and J.K., who were residents of a Japanese nephrology fellowship programme after the junior resident programme, completed the SAQ-NBR. They were doctors in their first, third, and fourth years of the programme, respectively. These three participants were asked to answer all the questions within 2 h. They were prohibited from searching any information related to the questions, such as through using the Internet or referring to references.

### Data Analysis

The overall correct answer rates for the five years and the correct answer rates by year were calculated for ChatGPT-3.5, ChatGPT-4, and Bard, and whether they met the pass criteria of a correct answer rate of ≥60% for each year was evaluated. The correct answer rates for each of the four categories–image, question type, taxonomy, and subspeciality– were calculated. Finally, differences between the scores of the nephrology residents were evaluated. Statistical analyses were performed using the R version 4.2.3. The Chi-square test or Fisher’s exact test was performed to compare each correct answer rate, and statistical significance was set at p < 0.05.

## RESULTS

### Performance by Overall Correct Answer Rates and Exam Year

Out of a total of 99 questions from 5 years of the SAQ-NBR, 61 were clinical and 38 were general questions. Among the clinical questions, 15 contained images. The overall correct answer rates were 31.3% (31/99) for ChatGPT-3.5, 54.5% (54/99) for ChatGPT-4, and 32.3% (32/99) for Bard. ChatGPT-4 had a significantly higher rate than both ChatGPT-3.5 and Bard (p = 0.002 and p = 0.003, respectively) (Table 1). Regarding passing standards for each year, neither ChatGPT-3.5 nor Bard met the criteria, whereas ChatGPT-4 met the criteria for 3 years.

**Table 1:** Correct answer rates of ChatGPT-3.5, ChatGPT-4, and Bard by exam year on the Self-Assessment Questions for Nephrology Board Renewal.

| Exam Year | Correct answer rates |  |  | p-value |  |  |
| --- | --- | --- | --- | --- | --- | --- |
|  | ChatGPT-3.5 | ChatGPT-4 | Bard | 3.5 vs 4 | 3.5 vs Bard | 4 vs Bard |
| 2018 | 8/20 (40.0%) | 12/20 (60.0%) | 9/20(45.0%) | 0.343 | 1.000 | 0.527 |
| 2019 | 4/20 (20.0%) | 13/20 (65.0%) | 6/20(30.0%) | 0.010 | 0.716 | 0.056 |
| 2020 | 5/19 (26.3%) | 7/19 (36.8%) | 3/19(15.8%) | 0.728 | 0.693 | 0.269 |
| 2021 | 6/20 (30.0%) | 10/20 (50.0%) | 4/20(20.0%) | 0.333 | 0.716 | 0.096 |
| 2022 | 8/20 (40.0%) | 12/20 (60.0%) | 10/20(50.0%) | 0.343 | 0.751 | 0.751 |
| Overall | 31/99 (31.3%) | 54/99 (54.5%) | 32/99(32.3%) | 0.002 | 1.000 | 0.003 |

Performance of ChatGPT and Bard on Self-Assessment Questions for Nephrology Board Renewal. Overall performance and exam year breakdown are reported. Differences in performance between large language models were queried using chi-squared and Fisher's exact tests.

### Performance by Four Categories

Regarding taxonomy, for the problem-solving questions, ChatGPT-4 had a significantly higher correct answer rate than ChatGPT-3.5 (64.0% vs. 28.0%; p = 0.022) and Bard (64.0% vs. 32.0%; p = 0.046) (Table 2). Regarding the question type, ChatGPT-4 had a significantly higher correct answer rate than ChatGPT-3.5 (57.8% vs. 28.9%; p = 0.020) and Bard (57.8% vs. 26.3%; p = 0.010) for clinical questions. In the image category, for the non-image questions, ChatGPT-4 had a significantly higher correct answer rate than ChatGPT-3.5 (58.3% vs. 34.5%, p = 0.003) and Bard (58.3% vs. 33.3%, p = 0.002). In the CKD subspeciality, ChatGPT-4 had a significantly higher correct answer rate than Bard (60.9% vs. 21.7%, p=0.016). No significant differences were observed in any of the other categories.

**Table 2:** Correct answer rates of ChatGPT-3.5, ChatGPT-4, and Bard by four categories on the Self-Assessment Questions for Nephrology Board Renewal.

| Category | Correct answer rates |  |  | p-value |  |  |
| --- | --- | --- | --- | --- | --- | --- |
|  | ChatGPT-3.5 | ChatGPT-4 | Bard | 3.5 vs 4 | 3.5 vs Bard | 4 vs Bard |
| **Image** |  |  |  |  |  |  |
| Non-Image Questions | 29/84 (34.5%) | 49/84 (58.3%) | 28/84 (33.3%) | 0.003 | 1.000 | 0.002 |
| Image Questions | 2/15 (13.3%) | 5/15 (33.3%) | 4/15 (26.7%) | 0.390 | 0.651 | 1.000 |
| **Question Type** |  |  |  |  |  |  |
| General Questions | 20/61 (32.7%) | 32/61 (52.4%) | 22/61 (36.1%) | 0.436 | 0.849 | 0.101 |
| Clinical Questions | 11/38 (28.9%) | 22/38 (57.8%) | 10/38 (26.3%) | 0.020 | 1.000 | 0.010 |
| **Taxonomy** |  |  |  |  |  |  |
| Recall | 21/59 (35.6%) | 31/59 (52.5%) | 21/59 (35.6%) | 0.095 | 1.000 | 0.095 |
| Interpretation | 3/15(20.0%) | 7/15(46.7%) | 3/15(20.0%) | 0.245 | 1.000 | 0.245 |
| Problem-Solving | 7/25(28.0%) | 16/25(64.0%) | 8/25(32.0%) | 0.022 | 1.000 | 0.046 |
| **Subspeciality** |  |  |  |  |  |  |
| CKD/ESKD | 10/23 (43.5%) | 14/23(60.9%) | 5/23 (21.7%) | 0.376 | 0.208 | 0.016 |
| AKI | 0/3(0.0%) | 0/3(0.0%) | 0/3(0.0%) | - | - | - |
| Glomerular Diseases | 8/28(28.6%) | 14/28(50.0%) | 12/28(42.9%) | 0.171 | 0.403 | 0.789 |
| Tubulointerstitial Diseases | 2/11(18.2%) | 6/11(54.5%) | 5/11(45.5%) | 0.183 | 0.362 | 1.000 |
| Hypertension/Vascular Diseases | 3/8(37.5%) | 3/8(37.5%) | 0/8(0.0%) | 1.000 | 0.200 | 0.200 |
| Water/Electrolytes/Acid-Base Disorder | 4/10(40.0%) | 8/10(80.0%) | 4/10(40.0%) | 0.170 | 1.000 | 0.170 |
| ADPKD/Urology | 1/3(33.3%) | 1/3(33.3%) | 2/3(66.7%) | 1.000 | 1.000 | 1.000 |
| Basic medicine | 3/13(23.1%) | 8/13(61.5%) | 4/13(30.8%) | 0.111 | 1.000 | 0.238 |

ChatGPT and Bard performance is reported for each category of Self-Assessment Questions for Nephrologist Board Renewal. Differences in performance between large language models were queried using chi-squared and Fisher's exact tests. CKD, chronic kidney disease; ESKD, end-stage kidney disease; AKI, acute kidney injury; ADPKD, autosomal dominant polycystic kidney disease.

### Comparison to Nephrology Resident’s Performance

The overall correct answer rates of residents in their first, third, and fourth years of the nephrology fellowship programme were 36.4% (36/99), 49.5% (49/99), and 67.7% (67/99), respectively (Table 3). Both ChatGPT-3.5 and Bard had lower overall correct answer rates than all the residents, whereas ChatGPT-4 had a correct answer rate equivalent to that of the third- and fourth-year residents. Regarding passing standards, the first-year resident did not meet the criteria for any year, the third-year resident met them at 1 year, and the fourth-year resident met them at 4 years. The proportion of ChatGPT-4 meeting the passing criteria was intermediate between the rates of the third- and fourth- year residents.

**Table 3:** Correct answer rates of nephrology fellowship programme residents by exam year on the Self-Assessment Questions for Nephrology Board Renewal.

| Exam Year | Correct answer rates |  |  |
| --- | --- | --- | --- |
|  | First -Year | Third-Year | Fourth-Year |
| 2018 | 7/20(35.0%) | 12/20(60.0%) | 16/20(80.0%) |
| 2019 | 6/20(30.0%) | 8/20(40.0%) | 11/20(55.0%) |
| 2020 | 7/19(36.8%) | 9/19(47.4%) | 14/19(73.7%) |
| 2021 | 8/20(40.0%) | 10/20(50.0%) | 13/20(65.0%) |
| 2022 | 8/20(40.0%) | 10/20(50.0%) | 13/20(65.0%) |
| Overall | 36/99(36.4%) | 49/99(49.5%) | 67/99(67.7%) |

## DISCUSSION

In this study, we evaluated the performance of multiple LLMs for medical examinations in nephrology and compared them with those of nephrology residents. ChatGPT-4 significantly outperformed ChatGPT-3.5 and Bard in terms of the overall correct answer rate, meeting the passing standards for nephrology board renewal requirements in 3 out of 5 years. ChatGPT-4 performed significantly better in questions requiring advanced interpretation, clinical questions, and non-image questions than ChatGPT-3.5 and Bard. While the overall correct answer rates of ChatGPT-3.5 and Bard were lower than that of first-year resident, ChatGPT-4 had a correct answer rate equivalent to that of the third- and fourth-year residents. Prior research has reported on the performance of LLMs regarding the mandatory skills and general knowledge of medical doctors, such as the Japanese national medical exam and USMLE [2,3,19]. To the best of our knowledge, this is the first report to compare the performances of multiple LLMs and human doctors in nephrology. Our results provide insights into the potential effectiveness and limitations of LLMs in nephrology.

In this study, ChatGPT-4 demonstrated a significantly higher overall correct answer rate than ChatGPT-3.5 and Bard. Previous reports have indicated that ChatGPT-4 demonstrated better performance than ChatGPT-3.5 in the USMLE and ophthalmology speciality exams [6,20]. Furthermore, in a written examination on neurosurgery, ChatGPT-4 outperformed Bard in all subspecialities [21]. In nephrology, which was verified in this study, the results were consistent with those of previous reports on the performance of other medical examinations. ChatGPT-4 was released four months after the release of ChatGPT-3.5, and a significant improvement in accuracy was observed in such a short period [22]. Thus, nephrologists need to stay updated on the emergence and updates of new LLMs, which could significantly differ in performance from previous versions.

ChatGPT-4 performed significantly better in questions requiring advanced interpretation, clinical questions, and non-image questions than ChatGPT-3.5 and Bard. Previous studies have indicated that ChatGPT-3.5 tends to be less accurate in questions requiring higher-level interpretation than other categories [12,23]. Moreover, ChatGPT-4 has a higher correct answer rate than ChatGPT-3.5 and Bard for questions requiring high-level inference [21]. For image questions, none of the LLMs used in this study could input image information, and the answers were based solely on the text of the question. Therefore, the superiority of ChatGPT-4 in non-image questions might represent its superiority in questions where all input information can be utilised, and might indicate differences in performance with other LLMs in overall nephrology. These results indicate an improvement in the high-level inference ability of LLMs in the field of nephrology. LLMs are trained to recognise patterns and relationships between words in the training data [1,24]. Their advanced reasoning ability is expected to continuously improve with an increasing amount of training data and the fine-tuning of medical tasks. This advanced interpretive ability may be beneficial in clinical settings, which are more complex than written tests and require multifaceted reasoning skills.

ChatGPT-4 consistently outperformed Bard and ChatGPT-3.5 across all subcategories in taxonomy, question type, and images. However, many of these comparisons did not yield statistically significant differences. This may be attributed to the limited number of test questions employed, which may have been insufficient to achieve the statistical power required to detect significant differences. With an increase in the number of test questions, the likelihood of observing statistically significant differences may increase.

This study had four limitations. First, all LLMs performed poorly on image questions because they could not input image information, potentially leading to a lower overall correct answer rate than the actual performance of the LLMs. Future iterations of GPT-4 are expected to process the image information [22], leading to improved accuracy. Second, the study was limited to Japanese participants. The performance of LLMs can vary depending on the input language [22], and multilanguage evaluations are desirable in the future. Third, question leakage was possible; as the SAQ-NBR questions from 2020 to 2022 are publicly available on the Internet, some questions may have already been included in the training data of the LLMs. Fourth, the validity of the correct answer rate for the residents who answered may be uncertain. The correct answer rates might not be representative of each year of training, and because the co-authors provided answers, various biases may have been included.

In this study, we evaluated the performances of ChatGPT and Bard in the field of nephrology. Currently, none of the LLMs can replace the clinical practice, care provision, and medical education of nephrologists; however, owing to model updates and technological innovations, rapid improvements may continue to occur in the future. To implement these improvements, nephrologists should keep up with the latest information. Further investigations are necessary to evaluate the effectiveness of these tools in clinical applications and medical education in the field of nephrology.

## CONCLUSIONS

In the SAQ-NBR, ChatGPT-4 outperformed ChatGPT-3.5 and Bard and met the passing criteria. Its performance fell between that of third- and fourth-year residents. ChatGPT-4 demonstrated superior inference skills compared with the other LLMs. This represents a potential application of LLMs in nephrology. Although LLMs could potentially become useful tools in medical education and clinical training, further research is necessary to evaluate their effectiveness.

## Data Availability

Due to the proprietary nature of the data used for this study (Self-Assessment Questions for Nephrology Board Renewal), the authors cannot post the raw data used for the analysis. However, the authors are able to share a part of the collected data (ex. large language model responses, etc.) on request to other researchers who have access to this exam.

## ACKNOWLEDGEMENTS

We would like to thank Editage for editing and reviewing this manuscript for English language.

## ETHICS APPROVAL AND CONSENT TO PARTICIPATE

An Ethical Approval Statement was not required for this manuscript since no specific patient information were included.

## AUTHORS’ CONTRIBUTIONS

R.N., D.I. and Y.S. participated in the writing of the paper.

Y.I., F.K. and J.K. participated in answering the exam.

R.N., Y.I., F.K., J.K., D.I., Y.S. participated in the approval of the final manuscript.

